# Contributing Factors to Reversed Willingness to Vaccinate after the COVID-19 Pandemic: Insights from a National Panel Survey

**DOI:** 10.1101/2025.08.06.25333101

**Authors:** Koh Oikawa, Michio Murakami, Sae Ochi, Mei Yamagata, Asako Miura

## Abstract

**Introduction:** Vaccination shows effectiveness at personal and community levels. To manage vaccine demand and supply, effectively promote vaccination, and plan future vaccination strategies, it is crucial to understand the change in the population’s willingness over time. Extreme changes in vaccination willingness can affect vaccine oversupply and shortage. Few longitudinal studies have focused on vaccination willingness after the COVID-19 pandemic. Therefore, we sought to explore temporal changes in COVID-19 vaccination willingness and identify associated variables during and after the pandemic.

**Methods:** A 4-year panel survey was conducted in Japan between January 2020 and March 2024. To evaluate changes in COVID-19 vaccine willingness and identify associated factors, we assessed vaccine hesitancy in September 2022 (i.e., previously vaccinated individuals who did not wish to receive future vaccinations) and vaccine consistency in March 2024 (i.e., previously vaccinated individuals who wished to receive future vaccinations and wished to receive vaccination in September 2022). Modified Poisson regression was performed to examine the predictors of vaccine hesitancy and consistency.

**Results:** Vaccination willingness declined from 51.0 % in September 2022 to 19.9 % in March 2024. Male sex, interest in COVID-19, risk perception, age, and agreement with government policies were negatively associated with vaccine hesitancy. Family experience of infection emerged as a borderline-significant negative factor in vaccine consistency.

**Discussion:** These results indicate that populations with younger populations, women, and people with low dread risk of perception for COVID-19 may have a low initial vaccination demand, which may decrease even after the pandemic ends. Vaccine demand may change within the same population. Therefore, it is crucial to reassess and adjust distribution strategies based on population characteristics.

**Conclusion:** It is essential to consider these associated factors and adjust the balance between vaccine supply and demand over time to maintain adequate coverage and prevent waste of resources.

## 1. Introduction

Vaccination against infectious diseases is effective at the individual level for reducing the risk of infection, disease severity, and death[1,2]. At the population level, vaccination helps alleviate pressure on healthcare systems by lowering the number of infections and severe cases and contributes to preventing infectious disease outbreaks through herd immunity[3].

To effectively promote vaccination, it is essential to understand the population’s demand for vaccination. Vaccine demand reflects an individual’s willingness to receive a vaccination. Within any population, there are groups that accept vaccination and others that present vaccine hesitancy. Vaccines vary in their administration schedules; some provide lifelong immunity with a single dose, and others require multiple doses depending on disease prevalence or persistence of immunity. In the context of emerging infectious diseases or those that cause repeated outbreaks, multiple vaccinations are often required to achieve herd immunity. However, vaccine hesitancy hinders the achievement of adequate population-level immunity. In 2019, the World Health Organization (WHO) identified vaccine hesitancy as one of the top ten global health threats[4,5]. Because vaccination is generally voluntary, it is essential for authorities to understand the balance between vaccine demand and supply to ensure a stable supply at the population level.

The importance of managing the balance between vaccine demand and supply has been demonstrated in previous studies. During the 2009 outbreak of the novel influenza virus in Japan, the government secured influenza vaccines. However, a rapid decline in the number of cases and a decrease in public interest in vaccination caused a decrease in vaccine demand and an oversupply of vaccines. Consequently, supply doses remained in storage, and some were abandoned[6]. This experience highlights the difficulties in balancing vaccine supply and demand during outbreaks of emerging infectious diseases. Additionally, it demonstrates that securing an adequate supply is insufficient; without accounting for the vaccination willingness of the target population, a substantial portion of the supply may be wasted.

In recent years, all countries, including Japan, experienced a global pandemic caused by the coronavirus disease 2019 (COVID-19), starting in early 2020. The WHO officially declared a pandemic on March 11, 2020[7]; by September of that year, the WHO recommended the prioritization of the vaccination of vulnerable groups, including older adults, people with underlying health conditions, pregnant women, healthcare workers, and socioeconomically disadvantaged populations. In addition, the WHO suggested booster vaccinations to improve immune defense, particularly for these at-risk groups[8]. The WHO set a goal of vaccinating 70 % of the global population by mid-2022 and officially ended the Public Health Emergency of International Concern (PHEIC) for COVID-19 on May 5, 2023[8,9]. Presently, the WHO recommends routine booster vaccinations for individuals who have never received a COVID-19 vaccine, older adults with comorbidities, immunocompromised individuals, pregnant women, and healthcare workers[10].

The Japanese government designated COVID-19 as a “designated infectious disease equivalent to category 2” under the Infectious Disease Control Law in February 2020, starting with mandatory case reporting. Following the WHO recommendations, Japan launched a vaccination program in February 2021. As part of the national policy, medical care for COVID-19, including vaccination, was provided free of charge, allowing all citizens to receive the vaccine at no cost. On May 8, 2023, the Japanese government reclassified COVID-19 as a Category 5 infectious disease, similar to seasonal influenza, and ended mandatory case reporting on the same day[11]. Simultaneously, vaccination, testing, and treatment have transitioned to fee-based provisions. At present, COVID-19 vaccination is recommended only for individuals aged 65 years and older, individuals aged 60–64 years who have cardiovascular, respiratory, or renal impairments that limit daily activities, or people are living with AIDS[11]. In total, more than 430 million doses of COVID-19 vaccine have been administered in Japan, with 83.8 % of the population having received at least one vaccination dose as of April, 2024[12,13].

The willingness to receive the COVID-19 vaccination during the pandemic depended on various factors, including demographics and sociopsychological characteristics[5,14,15]. Factors associated with low vaccination willingness include concerns about vaccine safety, fear of side effects[14,15], and employer-mandated vaccination[5]. Meanwhile, people expressed a higher vaccination willingness if they recognized the vaccine’s effectiveness[16], had already received the vaccine with their money, were encouraged to be vaccinated by healthcare providers or educational institutions, or had increased trust in vaccines following the COVID-19 pandemic[17]. According to a study conducted in Japan during the early phase of the COVID-19 pandemic, people with underlying health conditions, those who recognized the need for the vaccine, and those who wanted to protect others through their own vaccinations were more likely to demonstrate a high vaccination willingness [18]. Furthermore, longitudinal tracking of vaccination willingness revealed that a study conducted in 2021 identified a higher vaccination willingness when people perceived a sense of group responsibility to protect others[19]. In addition, a longitudinal study comparing vaccination willingness in 2021 with actual vaccination in 2022 identified factors contributing to the gap between past willingness and behavior. These factors included overall health status, history of annual influenza vaccination, prior COVID-19 infection, and engagement in preventive measures [20]. Meanwhile, changes in policies for COVID-19 affected vaccination willingness. In China, during the COVID-19 pandemic, the government implemented a strict “zero-COVID” policy, which included lockdowns and mobility restrictions. However, after the policy ended, people who experienced adverse effects following vaccination and had low trust in vaccines were found to have a lower vaccination willingness [21]. Therefore, extreme changes in vaccination willingness can affect both vaccine oversupply and shortage. However, most longitudinal studies on vaccination willingness have focused on the early phase of the COVID-19 pandemic and have extended only through early 2023. Few studies have investigated vaccination willingness after the WHO ended the PHEIC. A cross-sectional study conducted in Zambia in 2023 revealed that the recognition of COVID-19 severity, educational history, household income, and sex were associated with vaccination willingness [22]. However, further research, including longitudinal studies, is required to clarify the factors affecting vaccination willingness after the end of the pandemic.

Considering that Japan has administered over 300 million COVID-19 vaccine doses through multiple vaccinations[13], it is necessary to understand not only the initial vaccination willingness but also how such willingness may change after the end of the pandemic. Therefore, we sought to explore temporal changes in COVID-19 vaccination willingness and identify associated variables during and after the pandemic using a panel survey in Japan from January 2020 to March 2024, a period marked by drastic shifts in public health policies by the WHO and Japanese government.

## 2. Methods

### 2-1. Ethical approval

This study was approved by the Behavioral Subcommittee of the Research Ethics Committee of the Graduate School of Human Sciences, University of Osaka (authorization numbers: HB019-099: until February 2023; HB022-117: from February 2023 onwards). Informed consent was obtained online from all participants before the survey.

### 2-2. Participants and Recruitment

The participants were residents of Japan registered on the crowdsourcing platform, Crowd Works Inc. (Tokyo, Japan). As of July 2024, the platform had 6.72 million registered users aged ≥18 years [23]. Surveys were conducted between wave 1 (January 31 – February 1, 2020) and wave 30 (March 25 – 27, 2024), at intervals of approximately 2 weeks to 2 months[24]. Figure 1 shows the timeline of newly confirmed COVID-19 cases in Japan, national vaccination coverage, and the survey waves included in this study[13,25].

**Figure 1:**
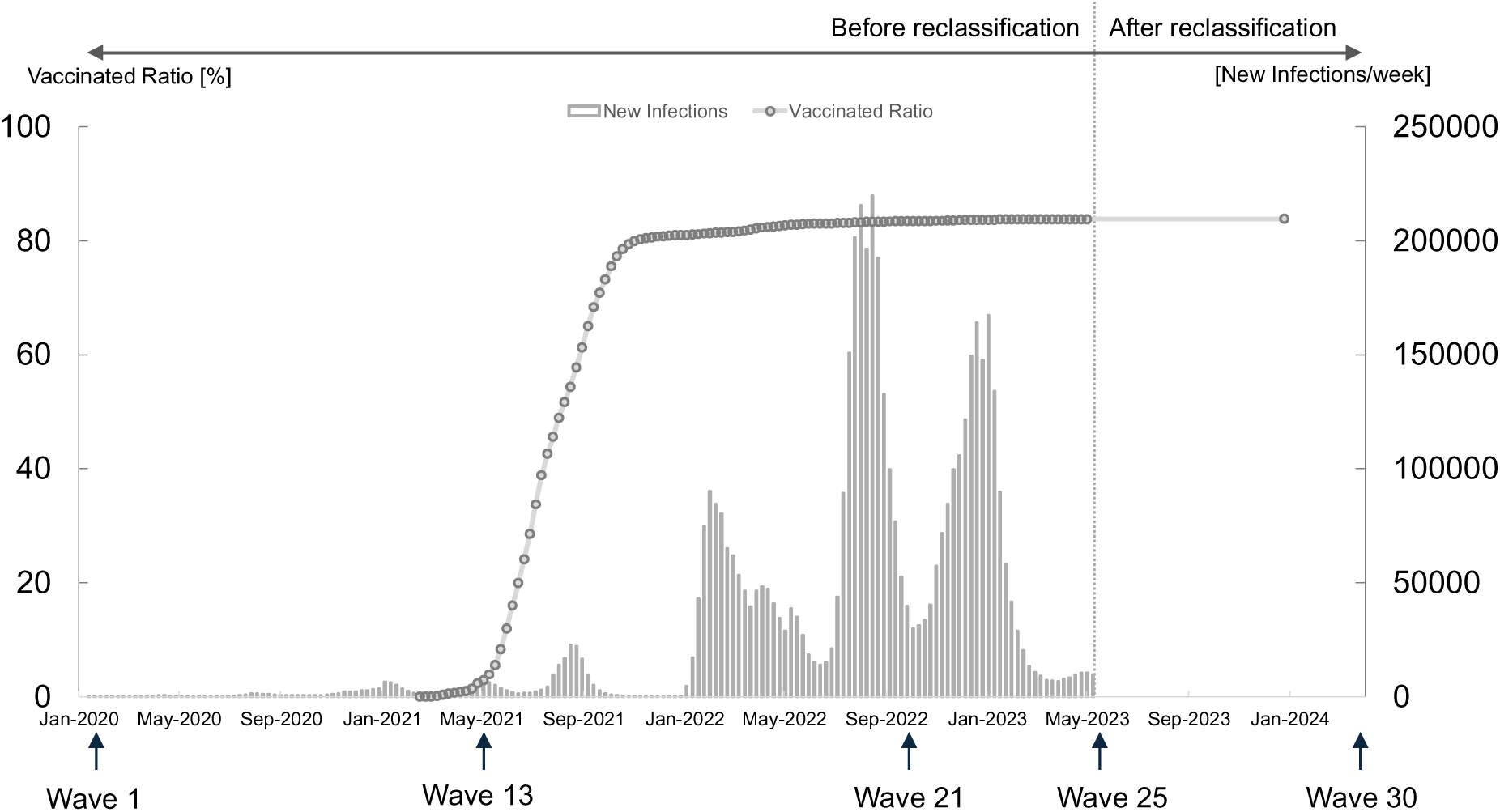
Timeline of newly confirmed COVID-19 cases in Japan[6,25], national vaccination coverage[13], and the survey waves included in this study. Following the reclassification of COVID-19 under the Infectious Disease Control Law in Japan in May 2023, mandatory nationwide case reporting ended; therefore, no data are available after May 2023.

To exclude participants who did not respond appropriately to the questions, we introduced the Directed Questions Scale (DQS), a previously reported method [26,27]. The DQS items were included in all survey waves, from wave 1 to wave 30. Participants who incorrectly answered two of the DQS items were excluded from the analysis.

In wave 1, 1,248 participants completed the survey. In wave 2, 1,200 of these original respondents were invited to participate in another survey. Subsequently, respondents in wave *N* were invited to wave *N*+1. In wave 13, 600 new participants were recruited. Accordingly, we defined respondents who had participated since wave 1 as “the original group” and those newly recruited in wave 13 as “the fresh group.” Participants received a reward of 120–150 Japanese yen for their participation in one survey.

Willingness to receive future vaccination was assessed beginning in wave 21 (September 27–29, 2022). In this study, we analyzed data from wave 21 and a final wave of 30. A total of 728 respondents completed wave 21, and 557 completed wave 30.

This study used a sample survey conducted in Japan. Based on a 95 % confidence interval and a 5 % margin of error, the minimum required sample size was 384[28]. Therefore, the number of respondents met the requirements for a statistically valid sample survey.

### 2-3. Questionnaire items

#### 2-3-1. Outcomes

In this survey, participants were asked about their COVID-19 vaccination status and willingness using five response options: “I have never been vaccinated and do not wish to be vaccinated in the future,” “I have never been vaccinated but willing to be vaccinated in the future,” “I have been vaccinated before but do not wish to be vaccinated in the future,” “I have been vaccinated before and willing to be vaccinated again,” and “Prefer not to answer.”

This study had two primary outcomes. First, vaccination willingness at wave 21 was assessed among participants who had previously been vaccinated. Responses indicating that individuals were not willing to receive future vaccination were defined as “vaccine hesitancy.” Second, vaccination willingness at wave 30 was assessed among those who had previously been vaccinated and were willing to vaccinate at wave 21. Responses indicating a willingness to receive future vaccinations were defined as “vaccine consistency.” Vaccine consistency was defined as the willingness to receive vaccine. The selection of vaccine consistency, instead of vaccine reverse willingness, as the outcome was based on the low extent of vaccine consistency. Hence, a result demonstrating a negative association with vaccine consistency is equivalent to a positive association with vaccine reverse willingness.

#### 2-3-2. Sociopsychological characteristics

Details of each panel item have been previously described[29–31]. We included the following sociopsychological characteristics: interest in COVID-19, agreement with government policy, dread risk perception, unknown risk perception, and pathogen avoidance tendency.

##### 2-3-2-1. Interest in COVID-19 and agreement with government policy

Interest in COVID-19 and agreement with government policies were included as sociopsychological variables, as their associations with willingness to receive COVID-19 vaccination have been previously reported [32,33].

##### 2-3-2-2. Risk perception

Slovic (1987) identified two types of risk perception–dread and unknown risk perceptions–as essential components of how people perceived risk[34]. Dread risk perception is characterized by factors such as uncontrollability, fear, potential for catastrophic outcomes, and lethality of consequences. Unknown risk perception is defined by attributes such as unobservability, unfamiliarity, novelty, and difficulty understanding exposure. Previous studies have revealed associations between risk perception and willingness to receive COVID-19 vaccination and preventive behaviors[29,35]. Dread and unknown risk perceptions were assessed using the mean score for each item.

##### 2-3-2-3. Pathogen avoidance tendency

The purity orientation–pollution avoidance scale, developed in Japan in 2021[36], allows for the measurement of the value of purity-based beliefs in the Japanese population. We included the five items in the scale for evaluate pathogen avoidance tendency because people with a stronger tendency to avoid pathogens engaged more frequently in preventive behaviors [37], and pathogen avoidance has been reported to be positively associated with willingness to receive COVID-19 vaccination[38]. The mean score of the five items was calculated for pathogen avoidance tendency.

We considered the responses in wave 21 for COVID-19 interest, dread risk perception, unknown risk perception, and pathogen avoidance tendency. Regarding agreement with government policy, we used the responses from both waves 21 and 25 (May 24–26, 2023). Wave 25 was selected because it was the closest survey period to the reclassification of COVID-19 to category 5 under Japan’s infectious disease control law on May 8, 2023, a point marked by a drastic policy shift.

##### 2-3-2-4. Information exposure

The informational panel items included medical information exposure (official websites of the Ministry of Health, Labour, and Welfare and medical institutions), mass media exposure, social networking service (SNS) exposure, and information exchange with acquaintances. These items were included based on previous studies that reported associations between information-seeking behaviors and engagement in preventive actions against COVID-19[30,39]. Because all these items were assessed in wave 2, responses from wave 2 were used for the analysis. We used a 7-point Likert scale for the following items: COVID-19 interest, agreement with government policy, dread and unknown risk perceptions, pathogen avoidance tendency, medical information exposure, mass media exposure, SNS exposure, and information exchange with acquaintances.

##### 2-3-2-5. Knowledge

Knowledge-related items included scientific knowledge and knowledge of COVID-19 reclassification. Vaccine-related knowledge is associated with vaccination willingness and treatment-related behaviors[39,40]. Therefore, these items were included in this study. The scientific knowledge items consisted of questions assessing basic biological (e.g., “Antibiotics kill viruses as well as bacteria”) and general scientific knowledge (e.g., “Early humans lived with dinosaurs during the same period.”). The COVID-19 reclassification knowledge items assessed the knowledge of the participants regarding changes associated with Japan’s reclassification of COVID-19 to category 5. Details of these knowledge items are described in a previous study[31]. Responses from wave 3 were used for scientific knowledge, as this item was assessed in that wave, while responses from Wave 25—responses provided at a period closest to the reclassification of COVID-19 to category 5—were used for COVID-19 reclassification knowledge. For both items, we used the number of correct answers to the five questions as the score.

##### 2-3-2-6. Infection experience

Regarding the COVID-19 infection-related individual items, we used responses regarding infection experience (personal/family) and incidence (personal/family). The experience of COVID-19 can affect subsequent vaccination willingness [41]. Given that the study outcome was vaccination willingness at waves 21 and 30, we defined infection experience (personal/family) as a previous COVID-19 infection in wave 21 and infection incidence (personal/family) as without COVID-19 infection in wave 21 but infection in wave 30.

##### 2-3-2-7. Background characteristics

Individual sociodemographic variables included sex, age, residential area, and occupation. These items were selected based on previous studies reporting their associations with the willingness to receive the COVID-19 vaccination [5,14,15,20,33]. Sex, age, and residential area were assessed in wave 1 for the original group and in wave 13 for the fresh group.

Regarding residential areas, the 15 prefectures containing Tokyo and government-designated cities were categorized as urban, whereas the remaining 31 prefectures were classified as non-urban. Occupation was assessed in wave 4 and categorized into three groups: (1) company employees and similar occupations, (2) self-employed, and (3) others.

### 2-4. Statistical analysis

Among the respondents in wave 21, we excluded those who did not report their sex, did not respond to the question on willingness to receive vaccination, or had never been vaccinated. The remaining respondents were classified into participant group A.

Afterward, among participant group A, those who expressed vaccination willingness in wave 21 and then responded to the survey in wave 30 were defined as participant group B. The numbers of respondents before and after exclusion are shown in Figure 2.

**Figure 2.**
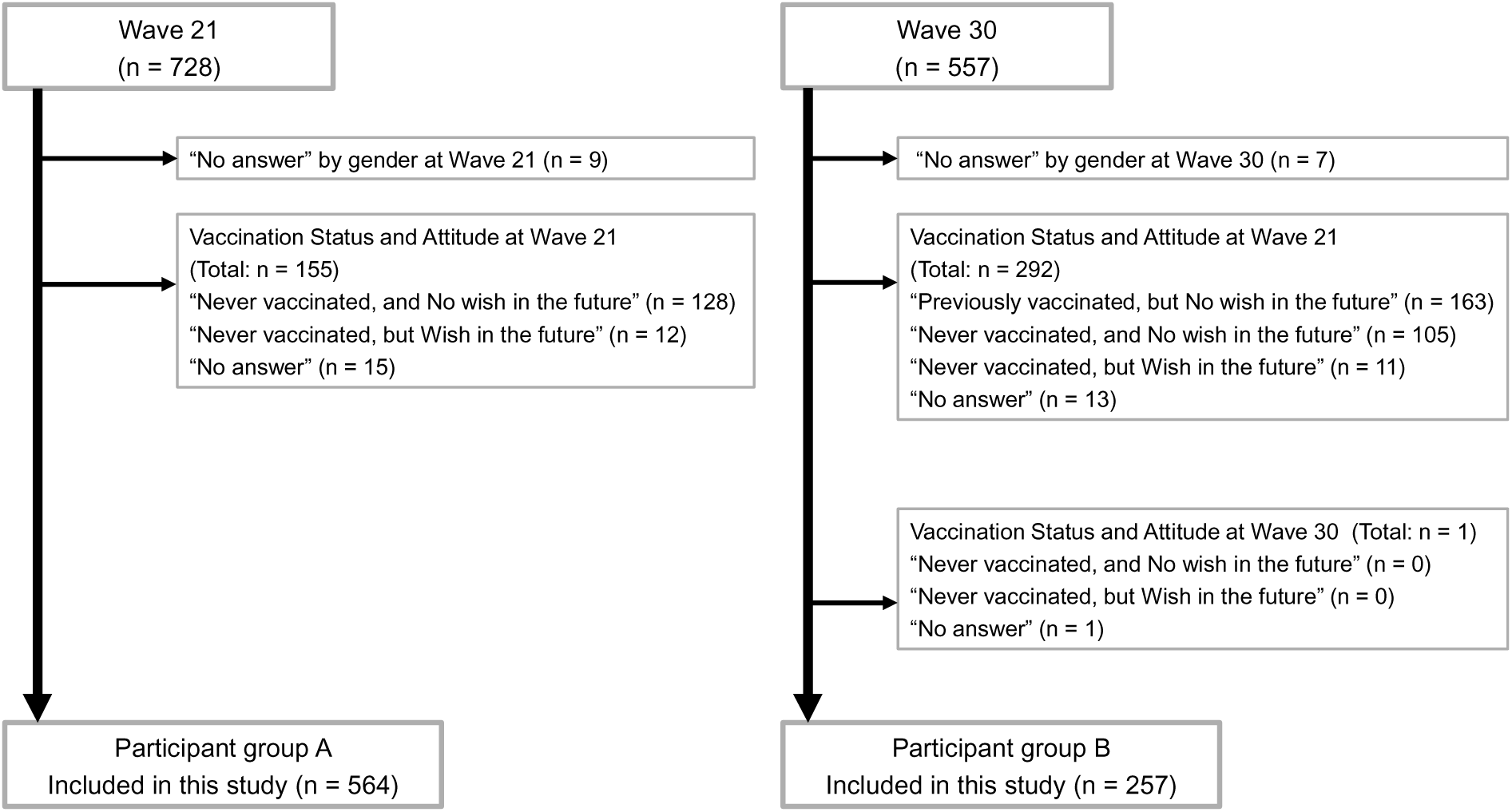
Number of respondents before and after exclusion.

Of the 728 individuals who participated in wave 21, 564 (77.5 %) were included in participant group A. Of the 557 respondents in wave 30, 257 (46.1 %) were included in participant group B.

First, univariate analysis using chi-square tests and t-tests was conducted to examine the relationships between each independent variable and two outcomes (vaccine hesitancy at wave 21 and vaccine consistency at wave 30) in participant groups A and B. Four items were included in the analysis for vaccine consistency at wave 30—infection incidence (personal and family), agreement with government policy at wave 25, and COVID-19 reclassification knowledge—because these items were assessed after wave 21.

Variables with a *P*-value < 0.1 in univariate analysis were included in multivariable analysis. A modified Poisson regression was performed for multivariable analysis. Variance inflation factors (VIFs) were calculated to detect multicollinearity.

For the 564 respondents included the new group in wave 13, the survey responses did not exist for the following variables: scientific knowledge, medical information exposure, mass media exposure, SNS exposure, information exchange with acquaintances, and occupation. Therefore, based on the inclusion of any of these items in the multivariable models, the analysis was divided into three parts: Analysis A-1 excluded the fresh group and included the above items; Analysis A-2 included the fresh group and excluded the above items; and Analysis B focused on participant group B.

There were no missing data for any items used in this analysis.

All analyses were performed using SPSS Ver. 28 (IBM Corp., Armonk, NY, USA), R version 4.4.2 (2024-10-31[42]) and rqlm package (Modified Poisson and Least-Squares Regressions for Binary Outcome and Their Generalizations; R package version 2.3-1[43]), and JMP® Pro 17.2.0 (JMP Statistical Discovery LLC, Cary, NC, USA).

## 3. Results

### 3-1. Vaccination willingness and changes

Among the 728 respondents in wave 21, nine who answered “no answer” for sex and 15 who responded “no answer” for vaccination willingness were excluded, and the remaining 704 were included as valid respondents. Of these, 347 had been vaccinated and were willing to be vaccinated again; 217 had been vaccinated but were not willing to be vaccinated again; 12 had never been vaccinated but were willing to be vaccinated; and 128 had never been vaccinated and were not willing to be vaccinated (vaccination rate: 80.1 %; proportion of those willing to be vaccinated in the future: 51.0 %). Similarly, of the 557 respondents in wave 30, seven answered “no answer” for sex and 12 responded “no answer” for willingness to vaccinate; the remaining 538 were included as valid respondents. Of these, 101 had been vaccinated and were willing to be vaccinated again, 325 had been vaccinated but were not willing to be vaccinated again, six had never been vaccinated but were willing to be vaccinated, and 106 had never been vaccinated and were not willing to be vaccinated (proportion of those willing to be vaccinated in the future: 19.9 %).

Table 1 presents the characteristics of the participants. The mean age was 42.6 years in participant group A and 44.7 years in participant group B. Regarding sex distribution, 57.4 % of the participants in group A were women and 42.6 % were men; in participant group B, 50.6 % were women and 49.4 % men.

**Table 1.**
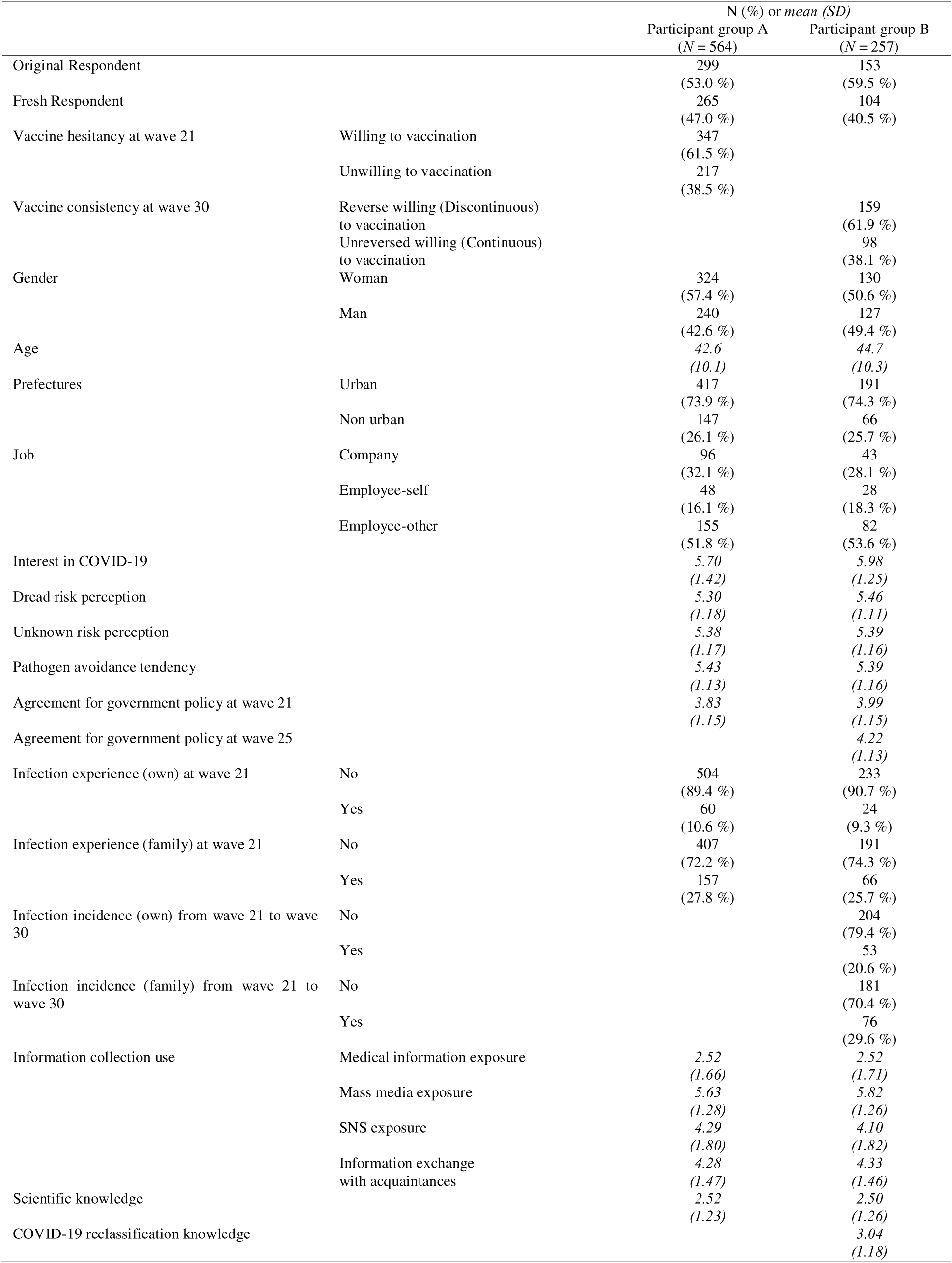
Participant characteristics.

In participant group A, 61.5 % responded willingness to vaccination (i.e., 38.5 % showed vaccine hesitancy). In participant group B, 61.9 % expressed vaccination willingness in wave 21 but no willingness by wave 30 (i.e., 38.1 % showed vaccine consistency).

### 3-2. Relationship between vaccine willingness and individual characteristics in univariate analysis

In participant group A, the following variables were positively associated with vaccine hesitancy: personal or family COVID-19 infection experience, lower interest in COVID-19, lower agreement with government policy, lower dread risk perception, smaller exposure to mass media, smaller exposure to information exchange with acquaintances, younger age, and female sex (Tables 2 and 3).

**Table 2.** Relationship between vaccine willingness and individual characteristics: Chi-square tests.

|  |  | Participant group A |  |  |  | Participant group B |  |  |  |
| --- | --- | --- | --- | --- | --- | --- | --- | --- | --- |
| | | N (%) | | P | $\phi$ or V | N (%) | | P | $\phi$ or V |
|  |  | Willing to vaccination | Unwilling to vaccination |  |  | Reverse willing (Discontinuous) to vaccination | Unreversed willing (Continuous) to vaccination |  |  |
| Gender | Woman | 189 (58.3%) | 135 (41.7%) | 0.070 | −0.076 | 89 (68.5%) | 41 (31.5%) | 0.028 | 0.137 |
|  | Man | 158 (65.8%) | 82 (34.2%) |  |  | 70 (55.1%) | 57 (44.9%) |  |  |
| Prefectures | Non-urban | 93 (63.3%) | 54 (36.7%) | 0.614 | 0.021 | 39 (59.1%) | 27 (40.9%) | 0.590 | −0.034 |
|  | Urban | 254 (60.9%) | 163 (39.1%) |  |  | 120 (62.8%) | 71 (37.2%) |  |  |
| Job | Company employee | 55 (57.3%) | 41 (42.7%) | 0.127 | 0.117 | 30 (69.8%) | 13 (30.2%) | 0.334 | 0.120 |
|  | Self-employed | 35 (72.9%) | 13 (27.1%) |  |  | 15 (53.6%) | 13 (46.4%) |  |  |
|  | Other | 104 (67.1%) | 51 (32.9%) |  |  | 55 (67.1%) | 27 (32.9%) |  |  |
| Infection experience (own) at wave 21 | No | 316 (62.7%) | 188 (38.3%) | 0.097 | 0.070 | 141 (60.5%) | 92 (39.5%) | 0.164 | −0.087 |
|  | Yes | 31 (51.7%) | 29 (48.3%) |  |  | 18 (75.0%) | 6 (25.0%) |  |  |
| Infection experience (family) at wave 21 | No | 260 (63.9%) | 147 (36.1%) | 0.064 | 0.078 | 111 (58.1%) | 80 (41.9%) | 0.035 | −0.131 |
|  | Yes | 87 (55.4%) | 70 (44.6%) |  |  | 48 (72.7%) | 18 (27.3%) |  |  |
| Infection incidence (own) from wave 21 to wave 30 | No |  |  |  |  | 122 (59.8%) | 82 (40.2%) | 0.181 | −0.083 |
|  | Yes |  |  |  |  | 37 (69.8%) | 16 (30.2%) |  |  |
| Infection incidence (family) from wave 21 to wave 30 | No |  |  |  |  | 114 (63.0%) | 67 (37.0%) | 0.570 | 0.035 |
|  | Yes |  |  |  |  | 45 (59.2%) | 31 (40.8%) |  |  |

**Table 3.** Relationship between vaccine willingness and individual characteristics: t-tests. SD: standard deviation.

|  | Participant group A |  |  |  |  | Participant group B |  |  |  |  |
| --- | --- | --- | --- | --- | --- | --- | --- | --- | --- | --- |
|  | *N* | Mean (SD) |  | *P* | *d* | *N* | Mean (SD) |  | *P* | *d* |
|  |  | Willing to vaccination | Unwilling to vaccination |  |  |  | Reverse willing (Discontinuous) to vaccination | Unreversed willing (Continuous) to vaccination |  |  |
| Age | 564 | 43.86 (10.45) | 40.59 (9.20) | <0.001 | 0.327 | 257 | 43.50 (9.72) | 46.56 (10.92) | 0.020 | −0.301 |
| Interest in COVID-19 | 564 | 6.02 (1.23) | 5.20 (1.56) | <0.001 | 0.601 | 257 | 5.89 (1.29) | 6.12 (1.18) | 0.153 | −0.184 |
| Dread risk perception | 564 | 5.48 (1.09) | 5.01 (1.25) | <0.001 | 0.404 | 257 | 5.36 (1.12) | 5.62 (1.10) | 0.074 | −0.230 |
| Unknown risk perception | 564 | 5.42 (1.17) | 5.30 (1.17) | 0.245 | 0.101 | 257 | 5.35 (1.16) | 5.45 (1.16) | 0.481 | −0.091 |
| Pathogen avoidance tendency | 564 | 5.41 (1.16) | 5.47 (1.09) | 0.500 | −0.058 | 257 | 5.30 (1.17) | 5.53 (1.14) | 0.124 | −0.198 |
| Agreement of policy at wave 21 | 564 | 3.98 (1.15) | 3.59 (1.10) | <0.001 | 0.351 | 257 | 4.01 (1.08) | 3.97 (1.26) | 0.803 | 0.032 |
| Agreement of policy at wave 25 |  |  |  |  |  | 257 | 4.30 (1.09) | 4.09 (1.19) | 0.149 | 0.186 |
| Medical information exposure | 299 | 2.59 (1.71) | 2.39 (1.57) | 0.315 | 0.122 | 153 | 2.43 (1.76) | 2.70 (1.60) | 0.357 | −0.157 |
| Mass media exposure | 299 | 5.78 (1.22) | 5.35 (1.34) | 0.005 | 0.342 | 153 | 5.85 (1.27) | 5.75 (1.24) | 0.657 | 0.075 |
| SNS exposure | 299 | 4.29 (1.80) | 4.30 (1.80) | 0.976 | −0.004 | 153 | 4.03 (1.79) | 4.25 (1.88) | 0.488 | −0.118 |
| Information exchange with acquaintances | 299 | 4.40 (1.41) | 4.05 (1.57) | 0.054 | 0.242 | 153 | 4.41 (1.36) | 4.19 (1.64) | 0.374 | 0.151 |
| Scientific knowledge | 299 | 2.49 (1.27) | 2.57 (1.15) | 0.584 | −0.066 | 153 | 2.42 (1.21) | 2.64 (1.35) | 0.301 | −0.176 |
| COVID-19 reclassification knowledge |  |  |  |  |  | 257 | 2.95 (1.24) | 3.17 (1.07) | 0.140 | −0.190 |

In participant group B, vaccine consistency was positively associated with a lack of family history of COVID-19 infection, higher dread risk perception, older age, and male sex.

### 3-3. Relationship between vaccine willingness and individual characteristics in multivariable analysis

In Analysis A-1, the following variables were significantly associated with vaccine hesitancy: interest in COVID-19 (incidence rate ratio [IRR]: 0.87, 95 % confidence interval [CI]: 0.78–0.97, *P* = 0.016) and agreement with government policy (IRR: 0.82, 95 % CI: 0.73–0.92, *P* < 0.001) (Table 4). Border significances were found for men (IRR: 0.76, 95 % CI: 0.55–1.04, *P* = 0.082), age (per 10 years) (IRR: 0.85, 95 % CI: 0.71–1.00, *P* = 0.051), and dread risk perception (IRR: 0.89, 95 % CI: 0.80–1.00, *P* = 0.052). Similar results were observed in Analysis A-2.

**Table 4.** Relationship between vaccine willingness and individual characteristics: modified Poisson regression. VIF: variance inflation factors, IRR: incidence rate ratio, CI: confidence Interval.

|  | Vaccine hesitancy |  |  | Vaccine hesitancy |  |  | Vaccine consistency |  |  |
| --- | --- | --- | --- | --- | --- | --- | --- | --- | --- |
|  | (Analysis A-1) |  |  | (Analysis A-2) |  |  | (Analysis B) |  |  |
|  | VIF | IRR (95%CI) | P | VIF | IRR (95%CI) | P | VIF | IRR (95%CI) | P |
| Gender (ref: woman) | 1.07 | 0.76<br>(0.55–1.04) | 0.082 | 1.05 | 0.80<br>(0.64–0.99) | 0.037 | 1.09 | 1.36<br>(0.99–1.86) | 0.057 |
| Age [per 10 years] | 1.11 | 0.85<br>(0.71–1.00) | 0.051 | 1.12 | 0.90<br>(0.81–1.00) | 0.046 | 1.04 | 1.16<br>(1.00–1.34) | 0.046 |
| Interest in COVID-19 | 1.60 | 0.87<br>(0.78–0.97) | 0.016 | 1.45 | 0.86<br>(0.80–0.92) | <0.001 |  |  |  |
| Dread risk perception | 1.35 | 0.89<br>(0.80–1.00) | 0.052 | 1.34 | 0.92<br>(0.85–1.00) | 0.055 | 1.03 | 1.17<br>(1.00–1.36) | 0.050 |
| Agreement of policy at wave 21 | 1.02 | 0.82<br>(0.73–0.92) | <0.001 | 1.01 | 0.85<br>(0.78–0.92) | <0.001 |  |  |  |
| Infection experience (own) at wave 21 (ref: No) | 1.20 | 1.20<br>(0.70–2.03) | 0.507 | 1.17 | 1.07<br>(0.78–1.47) | 0.689 |  |  |  |
| Infection experience (family) at wave 21 (ref: No) | 1.19 | 1.05<br>(0.74–1.51) | 0.775 | 1.17 | 1.09<br>(0.87–1.37) | 0.439 | 1.03 | 0.66<br>(0.44–1.01) | 0.056 |
| Mass media exposure | 1.15 | 0.94<br>(0.85–1.04) | 0.248 |  |  |  |  |  |  |
| Information exchange with acquaintances | 1.10 | 0.95<br>(0.86–1.05) | 0.293 |  |  |  |  |  |  |

In Analysis B, the following variables were significantly or border-significantly associated with vaccine consistency: men (IRR: 1.36, 95 % CI: 0.99–1.86, *P* = 0.057), age (per 10 years) (IRR: 1.16, 95 % CI: 1.00–1.34, *P* = 0.046), dread risk perception (IRR: 1.17, 95 % CI: 1.00–1.36, *P* = 0.050), and family COVID-19 infection experience (IRR: 0.66, 95 % CI: 0.44–1.01, *P* = 0.056).All VIFs were ≤ 1.60.

## 4. Discussion

We performed a longitudinal panel design study for 4 years, beginning at the onset of the COVID-19 pandemic. To the best of our knowledge, no previous studies have examined changes in the willingness to receive the COVID-19 vaccination and their contributing factors before and after the PHEIC.

### 4-1. Factors associated with vaccine willingness

The two main survey periods in this study were September 27–29, 2022 and March 25–27, 2024. As of September 2022, 80.1 % of the participants had been previously vaccinated. This number roughly matched with the national COVID-19 vaccination rate (83.4 %) in Japan at the same time[13], suggesting that our sample well represented the general population.

Furthermore, 61.9 % of respondents who had expressed willingness in September 2022 did not express vaccination willingness in March 2024.

Several studies have reported a reversal in vaccine willingness. A previous study in Japan between February–March 2021 and February–March 2022 (i.e., before and after the widespread of COVID-19 vaccination) reported that only 3.9 % of respondents who initially expressed willingness became unwilling to be vaccinated[20]. In the United States, an April and July 2021 study found that 17 % of the respondents who had previously expressed vaccination willingness later expressed vaccination unwillingness[44]. In China, where a strict zero-COVID policy was implemented, a March 2023 survey showed that 43.8 % of the respondents became unwilling to vaccinate after the policy ended[21]. In an October 2023 study in Zambia, performed after the end of PHEIC, approximately 37 % who had previously received one COVID-19 vaccination were unwilling to receive an additional dose[22].

Compared to these previous studies, our data showed a higher ratio of people who became unwilling to receive COVID-19 vaccination. Several factors at the time of our study may have contributed to this increased hesitancy. First, the predominant SARS-CoV-2 variant in Japan shifted from Delta to Omicron, which is generally associated with lower severity[45]. Second, the official recommendations for COVID-19 vaccination were reformed to narrow the target population[11]. Third, COVID-19 was reclassified as a category 5 infectious disease, which decreased the incentive for vaccination due to the introduction of fees[11]. These overlapping policies and virological and economic changes may have contributed to the increased rate of vaccination unwillingness in our study.

In Analyses A-1 and A-2, sex, interest in COVID-19, risk perception, age, and agreement with government policies were significantly associated with vaccination willingness. Besides, several studies have revealed factors associated with COVID-19 vaccination willingness. For example, an early 2021 study in the United Kingdom found that high COVID-19 interest was associated with a high vaccination willingness [32]. A multi country survey performed in seven European countries in April 2020 reported that younger women were less likely to be willing to receive the COVID-19 vaccine[15]. Moreover, other studies performed during the same period reported that women aged < 60 years were less willing to receive vaccinations [46,47]. In Japan, several studies conducted between 2020 and 2021 indicated that female sex and younger age were associated with lower vaccination willingness[18,48]. Risk perception was positively correlated with preventive health behaviors[35]. A literature review published between 2021 and 2022 revealed that inefficient government policies were negatively associated with vaccination willingness[47]. These earlier studies were conducted during the initial phase of the pandemic or around the early phase, when vaccines became available. Overall, our study, which was conducted in the later phase of the pandemic, showed similar associations between the vaccination willingness and factors such as sex, COVID-19 interest, risk perception, age, and agreement with government policy.

In contrast, our study did not find significant relationships between vaccination willingness and scientific knowledge, occupation, or personal/family experience with COVID-19. A previous study conducted in May 2020 among Japanese university students concluded that higher levels of basic scientific knowledge and educational history positively affected preventive behaviors against COVID-19[39]. In addition, a 2022 review indicated associations between past personal or family COVID-19 experience and vaccination willingness [47]. However, most studies were conducted during the initial phase of the pandemic. If the duration of the pandemic is prolonged, the increasing number of infections among acquaintances may have reduced vaccination willingness.

In Analysis B, participants were more likely to become unwilling to receive the COVID-19 vaccination if they had family infection experiences, were women, had a lower dread risk perception, or were younger. Notable, female sex, lower dread risk perception, and younger age were consistently associated with lower vaccination willingness in Analyses A and B. However, family infection experience was a borderline-significant factor in Analysis B, indicating a possible role in reversing vaccination willingness. In contrast, interest in COVID-19 and agreement with government policy were not associated with vaccination willingness in Analysis B.

Among previous studies performed during the pandemic, a 2020 U.S. study reported that individuals who were previously infected became more unwilling to receive vaccination[49]. In contrast, a 2021 study in Poland found that among unvaccinated people hospitalized due to COVID-19, those with severe symptoms expressed vaccination willingness [41]. These findings are consistent with those of previous studies. Acquired immunity through infection and lower severity of COVID-19 may have contributed to the decreased willingness to receive further vaccination. In addition, awareness of vaccination benefits may decline owing to family members’ experience of the infection.

### 4.2. Implications

Our findings revealed a huge decline in vaccination willingness (from 51.0 % in September 2022 to 19.9 % in March 2024); this decrease was observed over approximately 18 months.

Among younger populations, women, and people with a low dread risk of perception of COVID-19, not only may the initial demand for vaccination be low but the subsequent demand for vaccination in these groups may decline further as infections spread. In addition, because having a family member with COVID-19 infection was associated with a reduced vaccination willingness, vaccine demand at the population level may decrease as community-wide transmission increases.

Furthermore, neither interest in COVID-19 nor agreement with government policies was associated with changes in vaccination willingness. A high level of interest in COVID-19 or agreement with government policies does not explain the sustained willingness to receive vaccination over time. Therefore, even when interest in COVID-19 or agreement with government policy remains high, it is essential to recognize that vaccine demand may still decline in the long term, and vaccine supply must consider this potential reduction. In contrast, older adults continued to be highly willing to receive vaccination, both initially and after the infection became widespread. Because they are more susceptible to severe illness from infectious diseases and COVID-19 poses a high risk of severe outcomes in older adults, multiple vaccine doses have been recommended for this population[10,45]. Therefore, it is essential to consider vaccine supply to these high-risk groups.

When an infectious disease emerges and vaccines are ready to be distributed to the public, it is crucial to assess the population characteristics of the target group to determine how the prevalence of the variables identified in this study is associated with vaccination willingness. This assessment is essential to optimize the balance between vaccine demand and supply at different stages of the disease outbreak. Japan has a history of vaccine oversupply during the early stages of infectious disease outbreaks. During the novel influenza virus pandemic, a large quantity of vaccines was distributed in the early stages, resulting in abandoned stocks[6]. Similarly, in the case of COVID-19, approximately 240 million vaccine doses have been discarded in Japan[50]. Vaccine demand may change even within the same population, depending on public characteristics, particularly when comparing the initial and prolonged phases of an infectious disease outbreak. This implies that in the case of vaccines requiring multiple doses, such as the COVID-19 vaccine, it is crucial to reassess and adjust the distribution strategy based on the public characteristics of the target population when the outbreak is prolonged, even if the initial demand is adequately assessed. Modifying the vaccine distribution process according to population attributes is crucial to ensure delivery of vaccines in sufficient quantities not only to high-income countries but also to low- and middle-income countries, thereby avoiding wastage of limited vaccine resources.

### 4-3. Limitations

This study has several limitations. First, the participants in this study were registered users of a crowdsourcing platform, and their demographic characteristics may differ from those of the general Japanese population. However, given that rewards were provided to participants, even those who were not interested in the study were encouraged to participate, as noted in a previous study[30]. Moreover, the vaccination rate observed in our study closely matched the national vaccination rate in Japan, suggesting that the differences between our study population and the general population may be minimal.

Second, this study was performed in Japan, where public financial support for COVID-19 vaccination, differences in medical expenses, and the structure of the healthcare system may differ from those in other countries. Therefore, caution should be exercised when applying the findings of this study to other countries. Third, our panel survey did not reveal information about the severity of infection. A previous study reported that unvaccinated individuals who were hospitalized and required mechanical ventilation due to COVID-19 subsequently demonstrated an increased vaccination willingness [41]. In our study, it is possible that even among those with prior infection, differences in disease severity may have affected their vaccination willingness. The survey’s scope may need future expansion to include other nations’ insurance systems, vaccine policies, and the severity of COVID-19. Despite these limitations, our study provides relevant variables for changes in vaccination willingness, which should be considered in vaccine demand and supply.

## 5. Conclusion

In this study, we analyzed panel survey data collected between January 2020 and March 2024, focusing on responses from September 2022 and March 2024, to identify factors associated with willingness to receive the COVID-19 vaccine and changes in that willingness. During the 18-month survey period, willingness to receive the COVID-19 vaccine drastically declined from 51.0 % to 19.9 %. Female sex, younger age, and low dread risk perception were associated with vaccine hesitancy in September 2022 and with a reversed vaccination willingness in March 2024. Furthermore, family infection experience emerged as a borderline-significant predictive variable, suggesting its potential contribution to reversed vaccination willingness. Among populations with these characteristics, the willingness to vaccinate may change, especially when the vaccination phases are prolonged. To ensure an adequate and efficient supply of vaccines and avoid waste, it is essential to consider these associated factors and adjust the balance between vaccine demand and supply.

## CRediT Authorship statement

Koh OIKAWA: Conceptualization, Methodology, Formal analysis, Writing – Original Draft, Writing – Review & Editing. Michio MURAKAMI: Conceptualization, Methodology, Formal analysis, Writing – Review & Editing, Supervision, Project administration, Funding acquisition. Sae OCHI: Conceptualization, Methodology, Writing – Review & Editing, Supervision. Mei YAMAGATA: Conceptualization, Methodology, Investigation, Data curation, Writing – Review & Editing, Funding acquisition. Asako MIURA: Conceptualization, Methodology, Investigation, Data curation, Writing – Review & Editing, Funding acquisition.

All authors attest they meet the ICMJE criteria for authorship.

## Data availability

Data will be made available on request.

## Declaration of generative AI statement

During the preparation of this work, the authors used Chat GPT, DeepL, Ludwig, and Grammarly to search for articles and improve the grammar in the manuscript. Furthermore, the manuscript was edited by a professional native speaker. After using these services, the authors reviewed and edited the content as needed and took full responsibility for the content of the publication.

## Acknowledgements

We acknowledge the Editage for English language editing.

